# Exploring Polygenic Neuroimaging Derived Scores in a Longitudinal Attention-deficit/hyperactivity disorder Sample

**DOI:** 10.1101/2022.06.16.22276110

**Authors:** Tim van der Es, Sourena Soheili-Nezhad, Nina Roth Mota, Barbara Franke, Jan Buitelaar, Emma Sprooten

## Abstract

Genome-wide association studies (GWAS) indicate neuropsychiatric disorders to be highly polygenic. Polygenicity refers to the additive influence of multiple genes on variation in a disorder. GWAS have identified many single-nucleotide polymorphisms (SNPs) across the genome associated with neuropsychiatric disorders, each explaining a very small part of individual variance within a trait. This complicates the understanding of the genetic architecture and biological mechanisms underlying these disorders. Previous studies have successfully used common genetic variants associated psychiatric disorders to generate Polygenic Risk scores (PRS). PRSs estimate the aggregate genetic liability of an individual for a particular disorder or trait based on a genome-wide association study (GWAS) of said trait. Here, we present a novel bottom-up approach to polygenic scoring that starts at the brain, rather than at behavior or clinical diagnosis. We used GWAS of structural brain imaging derived phenotypes (IDPs) from the UK Biobank as a basis to generate polygenic imaging derived scores (PIDS). As a proof-of-concept of its application, we applied PIDS to quantify differences in the genetic influence on brain structure between persons with ADHD and unaffected controls. 94 IDPs were selected using the subcortical segmentation atlas and the Desikan-Killiany cortical atlas from FreeSurfer. In the polygenic model training stage, 72 out 94 PIDS were associated with their respective IDP in an independent sample. Global measures such as cerebellum white matter, cerebellum cortex and cerebral white matter ranked amongst the highest in variance explained ranging between 3% and 5.7%. Our results indicate that a majority of GWAS of structural neuroimaging traits are becoming sufficiently powered to enable reliable and meaningful use of polygenic scoring applications that accurately reflect the underlying polygenic architecture well. Larger discovery GWAS will further improve upon this. Conversely, our associations with ADHD were relatively weak. Larger target samples are required to establish robust links of PIDS with behavioral or clinical traits like ADHD. With this novel approach to polygenic risk scoring we provide a new tool for other researchers to build on in the field of psychiatric genetics.

## Introduction

Genome-wide association studies (GWAS) indicate neuropsychiatric disorders to be highly polygenic (Watanabe et al., 2019). Polygenicity refers to the additive influence of multiple genes on variation in a disorder. GWAS have identified many single-nucleotide polymorphisms (SNPs) across the genome associated with neuropsychiatric disorders, explaining a very small part of individual variance within a trait (Selzam et al., 2019; Liu et al., 2021). This complicates the understanding of the genetic architecture and biological mechanisms underlying these disorders. Genes are likely expressed in mediating brain development and information processing rather than influencing behavior directly (Weinberger, 2011). We will explore the use of imaging derived phenotypes (IDPs) to examine brain-based intermediate phenotypes, also known as endophenotypes. IDPs consist of structural measures of brain anatomy including tissue and structure volumes. We will use IDPs to assess the genetic architecture of neurological and psychiatric disorders and explain individual variation.

Human brain structure and function vary greatly between individuals. This variation can be measured non-invasively using magnetic resonance imaging (MRI) (Krain & Castellanos, 2006). The effects of neurological and psychiatric disorders such as Schizophrenia, Bipolar disorder, Autism and ADHD can be studied through MRI (Bonvicini et al., 2016; Romme et al., 2017; Yang et al., 2020). Understanding the genetic determinants of robust MRI markers could help identify biological pathways relevant to brain development and various diseases. Human brain structure traits possess attractive properties for genetic study (Matoba et al., 2020). Moreover, the study of human brain structure traits as broad endophenotypes for neuropsychiatric disorders could help in the identification of biological mechanisms and pathways that mediate individual differences in complex behaviors and vulnerability to disease (Bigos & Weinberger, 2010).

Given GWAS results, the count of risk alleles present in an individual person can be weighted by their effect size and summed into polygenic risk scores (PRSs). PRS is then used to quantify genetic liability for that individual. Mapping individual genetic liability can help understand the genetic architecture of the studied trait. Previously, studies have used hypothesis-driven approaches to investigating genetic associations with psychiatric disorders. A single PRS per neuropsychiatric disorder can fall short in describing variation for everyone due to biological heterogeneity. The genetic variation at the basis of risk factors may vary between individuals. With the availability of large data resources such as UK Biobank we can perform a hypothesis-free study. Here, we present a novel bottom-up approach to polygenic scoring that starts at the brain, rather than at behavior or clinical diagnosis. We used GWAS of structural brain IDPs from the UK Biobank as a basis to generate a set of polygenic imaging derived scores (PIDS) to better grasp the genetic variation between individuals.

ADHD will be used as a proof of concept for testing the potential utility of PIDS to clinical research. We chose ADHD because it is highly heritable with an average reported heritability of 76% (Bonvicini et al., 2016). Genetic factors have been broadly linked to the susceptibility to develop ADHD and some susceptibility genes have been identified (Demontis et al., 2022; Ronald et al., 2021). Differences in brain structure have been reported in cases with ADHD compared with controls (Krain & Castellanos, 2006). Alterations found in prefrontal gray matter, occipital gray matter, and inferior fronto-occipital fasciculus white matter were found not only in ADHD patients, but also in their unaffected siblings relative to controls (Bralten et al., 2016). Moreover, reductions in total cerebral volume, the prefrontal cortex, the basal ganglia (striatum), the dorsal anterior cingulate cortex, the corpus callosum and the cerebellum are also associated with ADHD (Poissant & Joyal, 2008; Curatolo et al., 2010).

In the present study we use a genomic approach to evaluate the genetic link between brain structure and disease risk. We will use PIDS to explore the relationship between ADHD and the brain, in a brain-driven fashion. The aim of this polygenic score was not to predict risk for individual patients but explore the construct and face validity of polygenic imaging derived scores. This new approach could improve our understanding of factors involved in neuropsychiatric traits.

## Methods

### Data

#### Discovery Sample

Summary statistics were obtained from the UK Biobank, a long-term epidemiological study of 500,000 volunteers from across the United Kingdom between the age of 40 and 69. The dataset used for this project contains 3929 IDPs using 39,691 participants and QC measures available from UKB ranging over six distinct MRI modalities. (Smith et al., 2021). For this project we focused on the structural image modality. After removal of individuals as part of genetic processing a sample of 33,224 individuals remained (*n* = 33,224, 17,411 genetic females). The ages in the sample were as follows: females mean age = 63.7±7.4 years (min = 45.1, max = 81.8); males mean age = 65.0±7.6 years (min 46.1, max 81.8). Additional information on the data can be found in the original paper by Smith et al., 2021. The UK Biobank has approval from the Northwest Multi-centre Research Ethics Committee (MREC).

#### Neuroimaging traits / IDPs

Freesurfer V6.0.0 (https://surfer.nmr.mghm.harvard.edu) was used to define both cortical and subcortical structures. The definition of cortical structures, referred to as parcellation, is performed by mapping atlas regions of the brain onto an individual’s cortical surface model (Destrieux et al., 2010). The Desikan-Killiany atlas was used for automated parcellation (aparc) of the cortical structures (Desikan et al., 2006).

For this project we were interested in broad structural differences of the brain between ADHD patients and controls. All gray matter tissue, global white matter and cerebrospinal fluid labels from Freesurfer were included in this project. These labels are part of the ‘regional and tissue volume category’ for 18 subcortical volumes from the subcortical segmentation atlas and 29 cortical volumes from the Desikan-Killiany atlas were included, bilaterally, resulting in 94 IDP’s (Table 1. Supplementary materials). Confounds were removed from the data including 40 population genetic principal components supplied by UKB and with a greatly expanded new set of confounds including age, head size, sex, head motion, scanner table position, imaging center and scan date-related slow drifts. More details for the imaging and genetics processing and cohorts can be found in the online methods of the original paper by Smith et al., 2021.

**Table 1.**
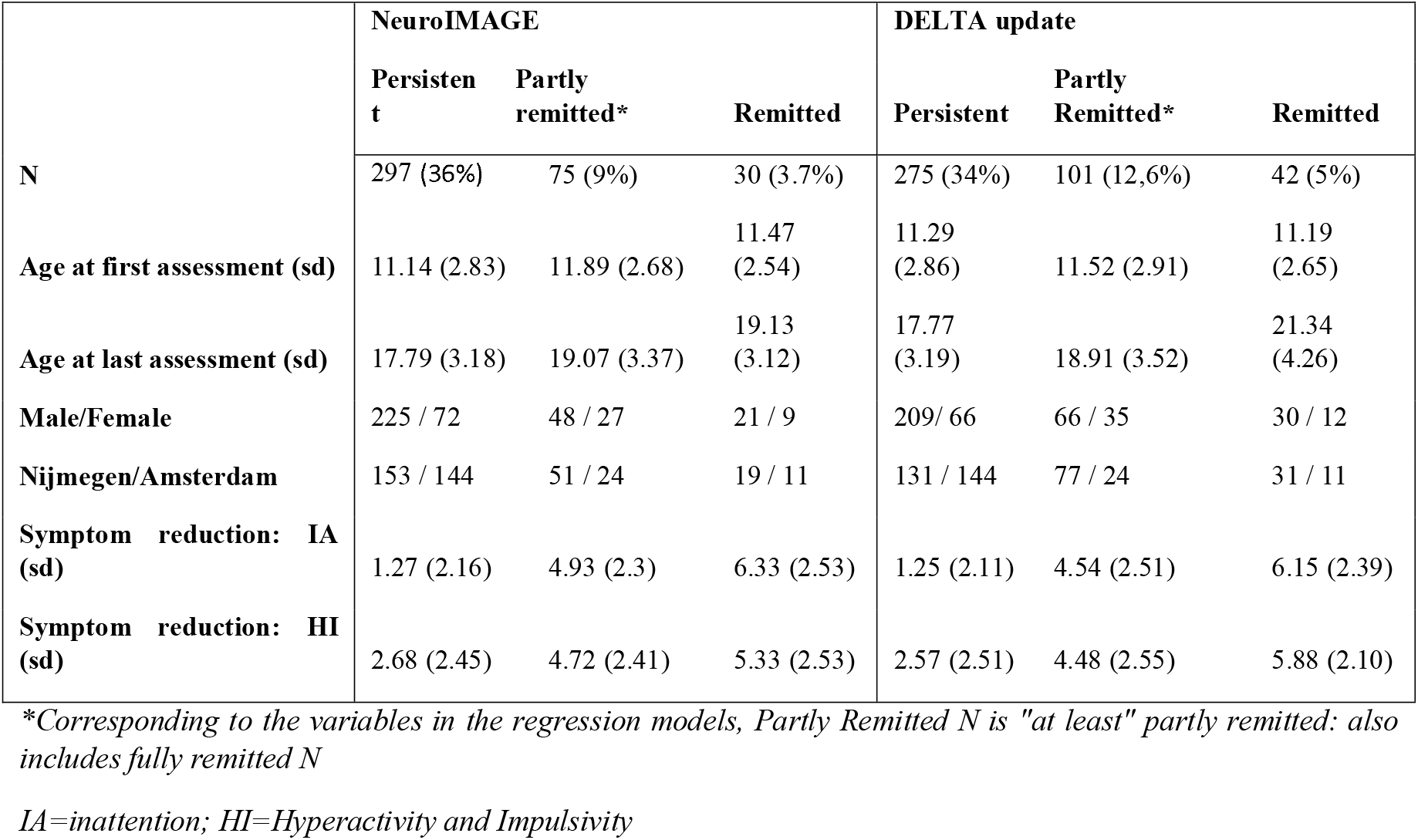
Demographic and clinical information of the NeuroIMAGE cohort per wave

#### Target Samples

The NeuroIMAGE study is a follow-up of the Dutch part of the International Multicenter ADHD Genetics (IMAGE) project (Müller et al., 2011). NeuroIMAGE sample is a longitudinal cohort of ADHD patients, their unaffected siblings, and controls. The original 365 ADHD families and 148 control IMAGE families, consisting of 506 participants with an ADHD diagnosis, 350 unaffected siblings, and 283 healthy controls (Kuntsi et al., 2006). In combination with new participants the NeuroIMAGE study consisted of 1,069 children (751 from ADHD families; 318 from control families) and 848 parents (582 from ADHD families, 266 from controls). The NeuroIMAGE dataset consists of 3 repeated measures ‘waves’, collected between 2003 and 2012. A slight adaptation of the Parental Account of Childhood Symptoms (PACS) clinical interview and the Conners Parent and teacher rating scales were used in wave 1 to more objectively asses children’s symptoms (Taylor, 1986; Conners, 1997,1998; Müller et al., 2011). In the immediate follow-up wave 2 & 3, Conners’ Adult ADHD Rating Scale (CARS R-L), Conners’ Parent Rating Scale (CPRS R:L), and Conners Wells’ Adolescent Self-Report scale: Short Form (CASS:S) (Conners et al., 1998; Conners et al., 1998; Parker & Sitarenios, 1999). MRI measurements were recorded for each child older than 7 years without contradiction for an MRI measurement (e.g., implanted metal and medical devices) and willingness. Freesurfer V6.0.0 (https://surfer.nmr.mghm.harvard.edu) was used to define both cortical and subcortical structures. The 94 IDP summary statistics from UKB were matched by volumetric measurements for the same cortical and sub-cortical brain regions. Participating children completed a session in a magnetic resonance imaging (MRI) scanner. Data was collected at the Radboud University Medical Centre, Donders Centre for Cognitive Neuroimaging or VU University Amsterdam and VU University Medical Centre. At both sites similar 1.5 T scanners were used (Siemens SONATA and Siemens AVANTO; Siemens, Erlangen Germany), both using 8-channel Phase Array Head Coils. The NeuroIMAGE study was approved by the regional ethics committee (Centrale Commissie Mensgebonden Onderzoek).

#### NeuroIMAGE

To determine ADHD diagnosis at the time of participation in NeuroIMAGE, a diagnostic algorithm was used combining the diagnostic interview (K-SADS) with the Conners rating scales as described previously in von Rhein et al (2015). Following the diagnostic algorithm of clinical interview and ADHD, participants were categorized as either diagnosed with ADHD, Unaffected, and ADHD Subthreshold. To be diagnosed with ADHD, participants were required to have ≥ 6 hyperactive/impulsive and/or inattentive symptoms, met the DSM-IV criteria for pervasiveness and impairment and showed an age of onset before 12. To be considered unaffected, participants were required to have a *T* < 63 on each subscale of the Conners questionnaires and ≤ 3 symptoms derived from the combined symptom counts of the K-SADS and CTRS-R: L/CAARS-S:L. Participants who did not meet the requirements for either ADHD or unaffected status received the ADHD-subthreshold classification. Criteria were adapted for adults (≥ 18 years) resulting in a diagnosis after a symptom count of ≥ 5. Adults were considered unaffected with an ADHD symptom count of ≤ 2. Table 1 summarizes the NeuroIMAGE sample according to clinical trajectory of persistence, partial remission, and full remission. Inclusion criteria for participants was a full diagnosis of ADHD at least once previously in NeuroIMAGE and having supplied quality-control passing DNA.

#### Target sample update DELTA

A subset of 60 NeuroIMAGE participants were reexamined in an additional MRI follow-up study, Determinants of Long-Term Trajectories of ADHD (DELTA), about five years after the third NeuroIMAGE wave. Participants were only included in DELTA if they had a full diagnosis of ADHD at least once previously in NeuroIMAGE and had contributed DNA which passed QC. The Diagnostic interview voor ADHD bij volwassenen (DIVA) interview was used to assess diagnosis of ADHD considering all participants were aged 18 or above (Kooij & Francken, 2010). DIVA is based on the DSM-IV criteria and is a follow up to earlier semi-structured interviews for ADHD in adults. Individuals were classified as “affected” if they had 6 or more symptoms for hyperactivity and/or inattention and clinically significant self-reported impairment in at least 2 domains. Individuals were labeled as “unaffected” with fewer than 3 symptoms for inattention and hyperactivity or “Subthreshold” if they fit neither category. Details concerning demographics and clinical information can be found in table 1. Individuals were categorized to be either (I) persistent, (II) partially in remittance or (III) fully remitted.

### Persistence vs Remittance

To define this longitudinal categorical status of participants, only the first and last measurement were considered in the NeuroIMAGE study. Individuals with a full diagnosis of ADHD at the first or second wave and at their last measurement were considered persisting. Individuals with a full diagnosis of ADHD at the first and/or second wave and were unaffected at the last available measurement were considered fully remitted. Individuals with a full diagnosis at the first or second wave and subthreshold ADHD or unaffected at the last measurement were considered (at least) in partial remission. Individuals only affected at the last measurement were considered ‘late onset’ and excluded (N=48). Details on the diagnostic tools, imaging and genetics processing and cohorts can be found in the original paper by van Rhein et al., 2015.

### Genotyping and quality control

DNA was extracted from whole blood or saliva. Genotyping was done using the Illumine PsyChip, in two batches (N=192 and N=798). Plink 1.9 was used to merge the batches ((https://www.cog-genomics.org/plink/1.9/) (Purcell, 2007; Yang et al., 2015). During quality control, mismatching SNPs between batches, sex chromosomes, insertions, deletions, other non-SNP variation; and ambiguous SNPs were excluded. RICOPILI pipeline was used to further process the data (https://sites.google.com/a/broadinstitute.org/ricopili). SNP exclusion criteria were set at call rate < 0 .05, Hardy-Weinberg equilibrium P,10^−6^, and minor allele frequency <0.01. Moreover, SNPs with a missing-rate difference > 0.01between batches were also excluded. Exclusion criteria for individuals were call rate < 0.05, identity by descent with another participant (quasi-random exclusion for each pair, with priority to more complete phenotyping data), inconsistent sex between self-report and GWAS data. 18 individuals were excluded in RICOPILI after principal component analysis (PCA) due to deviation from the main sample cluster (defined by 4 principal components, greater than 3 standard deviations). No association was found between the principal components and genotyping batch. A batch-effect of P,0.0008 resulted in the exclusion of 48 SNPs (P-threshold determined based on QQ-plot diagonal of the genome-wide batch effects). After quality control, 283,843 SNPs and 952 individuals were included in the re-run of the PCA analysis, required for subsequent analyses. Imputation was performed in accordance with the RICOPILI pipeline using minimac3 and Phase3 of the European 1000 genomes as reference. The PRS analysis were run with the strict best-guess output from RICOPILI (INFO score>0.1, MAF>0.005,2,840,886 SNPs).

### PIDS calculation

The PIDS analysis consisted of three different stages. (1) First, PRSice was run to determine optimal P-thresholds and select the model using IDP’s (Choi et al., 2020; Euesden et al., 2016). PIDS’s able to successfully predict their respective imaging phenotype were optimized. As we sought to validate the genomic signature of the PIDS, establishing a significant association between the PIDS and IDP in an independent sample verifies the predictive quality of the genetic score. (2) Secondly, the optimal model for each PIDS was used to predict ADHD diagnosis in the same dataset using generalized logistic mixed linear models to account for family structure. In addition, an explorative analysis was run to explore the consistency of associations between PIDS and ADHD during the different waves of NeuroIMAGE. (3) Thirdly, further analyses were performed using PIDs in trend association with ADHD diagnosis. General linear mixed effects models were constructed to regress the PIDs on persistence/remittance of ADHD symptoms in the NeuroIMAGE and DELTA dataset, accounting for family structure. ADHD-relevant PIDS were run on persistence and remittance in the longitudinal NeuroIMAGE cohort and consequent follow-up DELTA wave. An overview of all different steps performed in this project can be seen in figure 1.

**Figure 1.**
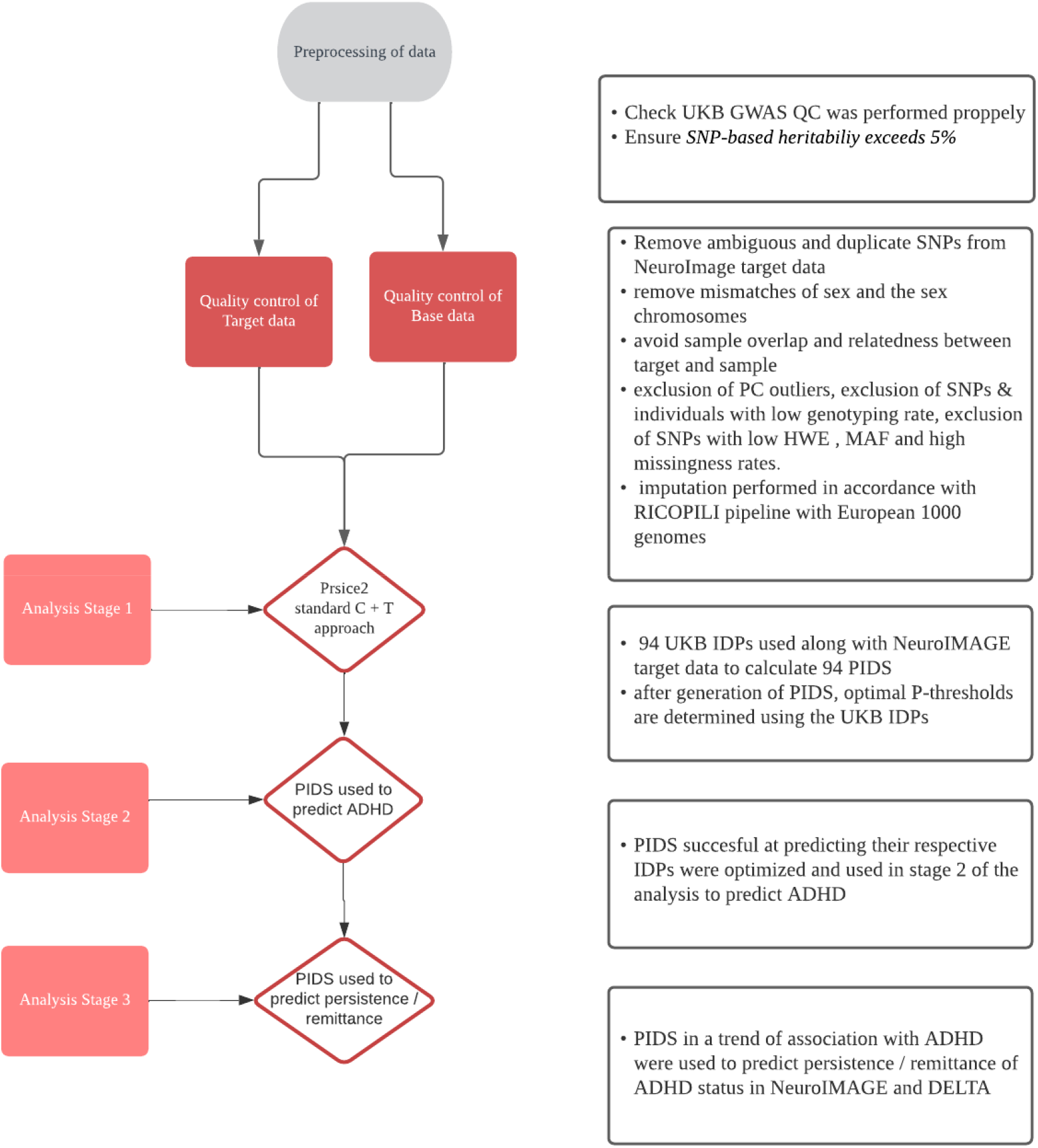
Flowchart of data quality control and analysis The project consisted of a data preprocessing stage and three stages of analysis. The software used for the calculation of the Polygenic Imaging Derived Scores (PIDS) is Prsice2.

Step 1: Model training and PIDS selection: The UKB summary statistics were used as the ‘base data’ PRSice input with standard clumping parameters (P=1, R^2^ <0.1 and 250kb interval) and P-value thresholds (P-value thresholds between P=10^-4 and P=0.5 at P=5*10^-5 intervals). We chose to run this broad selection of P-thresholds to thoroughly test the performance of the PIDS and see if the known polygenic nature of the IDPs was reflected across P-thresholds. In the resulting bar plots of variance explained across P-thresholds, we expect to see a broad range of multiple P-thresholds to perform well, including higher P-thresholds, reflecting the polygenicity of the IDPs. P-threshold optimization was based on the best high-resolution prediction of each PIDS on their respective IDP counterpart in the NeuroIMAGE sample before the update of DELTA. Because our aim was to establish whether an association exists between the PIDS and their respective IDP, for the PIDS selection, a lenient significance threshold of P,0.05 was chosen for PIDs selection in this training phase.

Step 2: Exploratory associations with ADHD. PIDS that passed the training stage at P<0.05 were entered in logistic mixed models run in R 3.6.1, with family as a random factor, ADHD diagnosis as dependent variable, PRS and as covariates, genotype batch, site and sex and four principal components. Afterwards, an exploratory analysis was performed in which the PIDS were regressed on ADHD and ADHD-subthreshold at waves one, two and three to ascertain whether any effects of the PIDS were consistent across time. Mixed logistic regressions were run for each PIDS found to be in association with ADHD following the exploratory analysis. For step 3, PIDS associated with ADHD were regressed on Persistence-Remittance, Partial, Remittance, and Remittance-Category from the NeuroIMAGE dataset as outcome variables. Four regression models were constructed for DIVA DELTA diagnosis (DSM5), DELTA diagnosis von Rhein et al. (DSM4), Persistence-Remittance-updated, and Partial Remittance-updated from the DELTA dataset (N=60).

### Power analysis

The sample included twin pairs and their siblings, meaning observations weren’t independent. Therefore, the effective sample size was calculated to perform the power analysis (N_E_ = (N*M) / (1 + ICC*(M-1)) in which N = the number of families, M= the number of individuals in a family and ICC = the (average) phenotypic correlation within a family. The power analysis itself was performed using the DubridgeLab library (https://rdrr.io/github/DudbridgeLab/avengeme/src/R/sampleSizeForGeneScore.R) based on the Power and Predictive Accuracy of Polygenic Risk Scores paper (Dudbridge, 2013). The PIDS were tested against ADHD to test for a common genetic basis with the Imaging Derived Phenotypes.

## Results

### Model optimization and calculation of PIDS

Descriptive information on each PIDS such as variance explained (*R*^*2*^), optimal P-threshold and SNP number can be found in the supplements (supplementary tables 1, 2 & 3). 72 out of 94 PIDSs were associated with their respective IDP (*p* < 0.05) in the target sample. The 72 PIDS were separated into 4 subgroups of left hemisphere cortical PIDS (1), right hemisphere cortical PIDS (2), left hemisphere Subcortical PIDS (3), and right hemisphere Subcortical PIDS (4). The genetic effect size (R^2^), P-threshold, and heritability (*h*^2^) for the PIDS in each subgroup are displayed in circosplots (figure 2-5).

**Figure 2.**
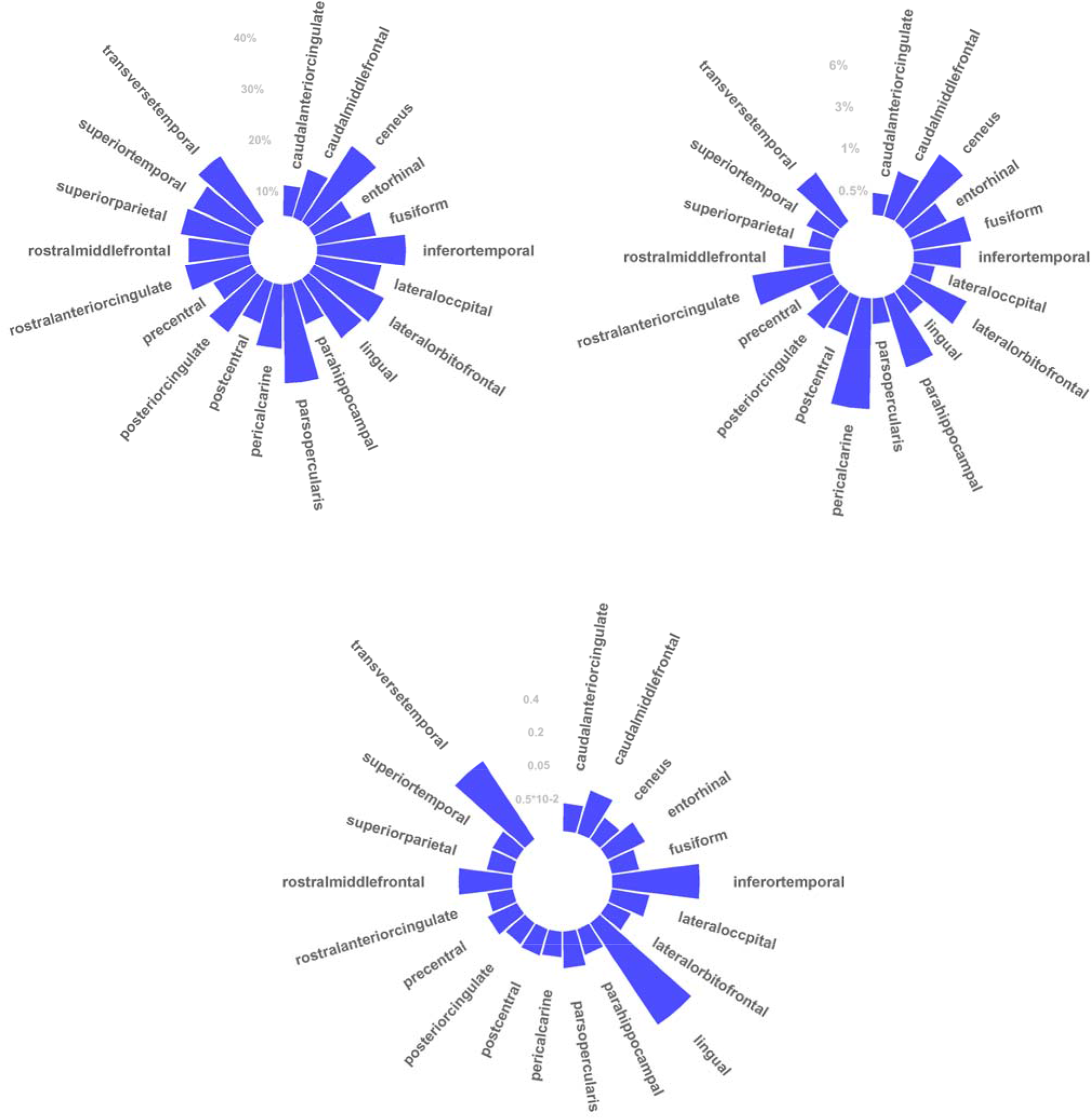
Circosplot of left hemisphere of the Cortical brain volume PIDS. Figure 2.1 circosplot of heritability, Figure 2.2 circosplot of variance explained, and Figure 2.3 circosplot of P-thresholds.

**Figure 3.**
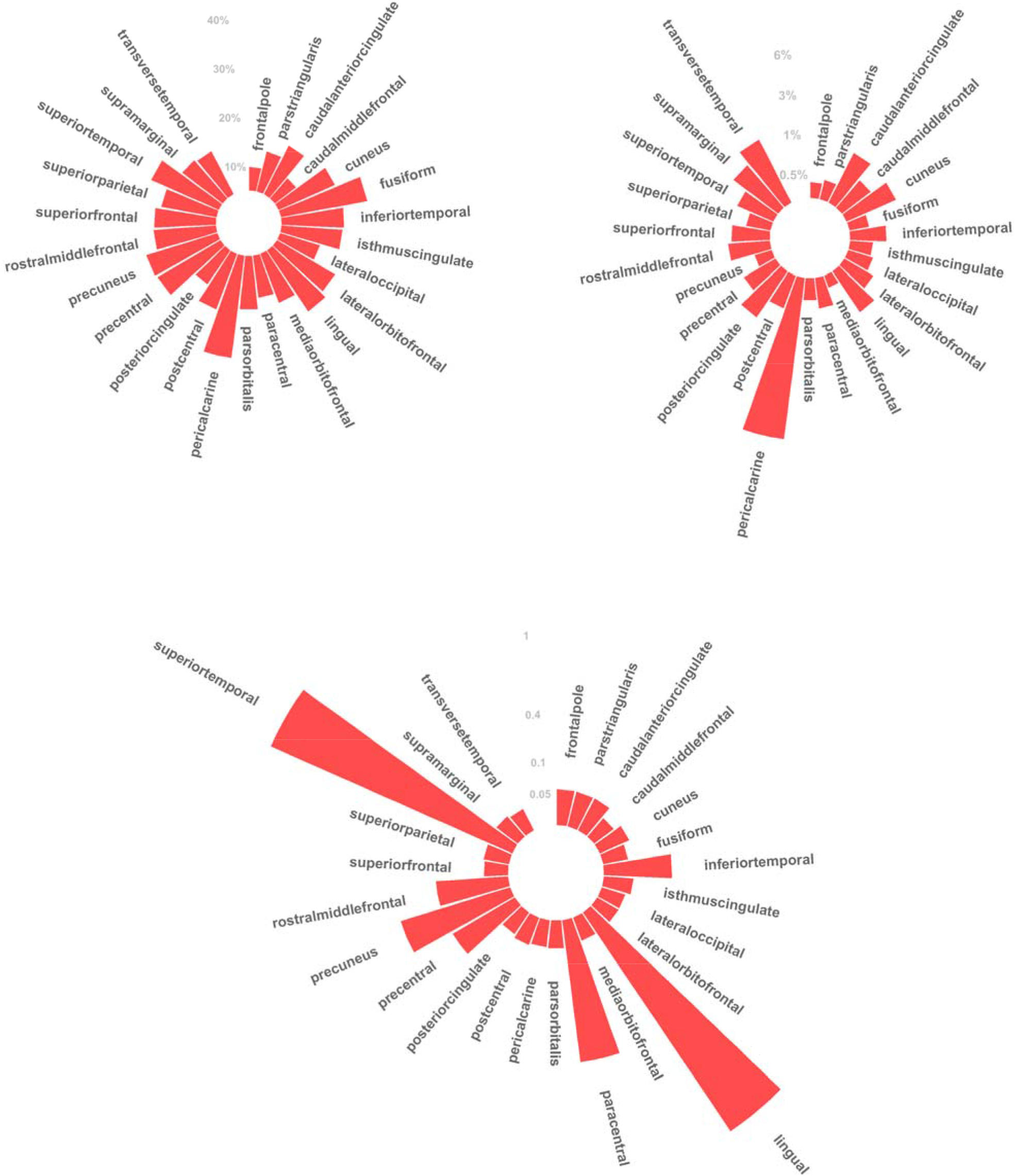
Circosplot of right hemisphere of the Cortical brain volume PIDS. Figure 3.1 circosplot of heritability, Figure 3.2 circosplot of variance explained, and Figure 3.3 circosplot of P-thresholds.

**Figure 4.**
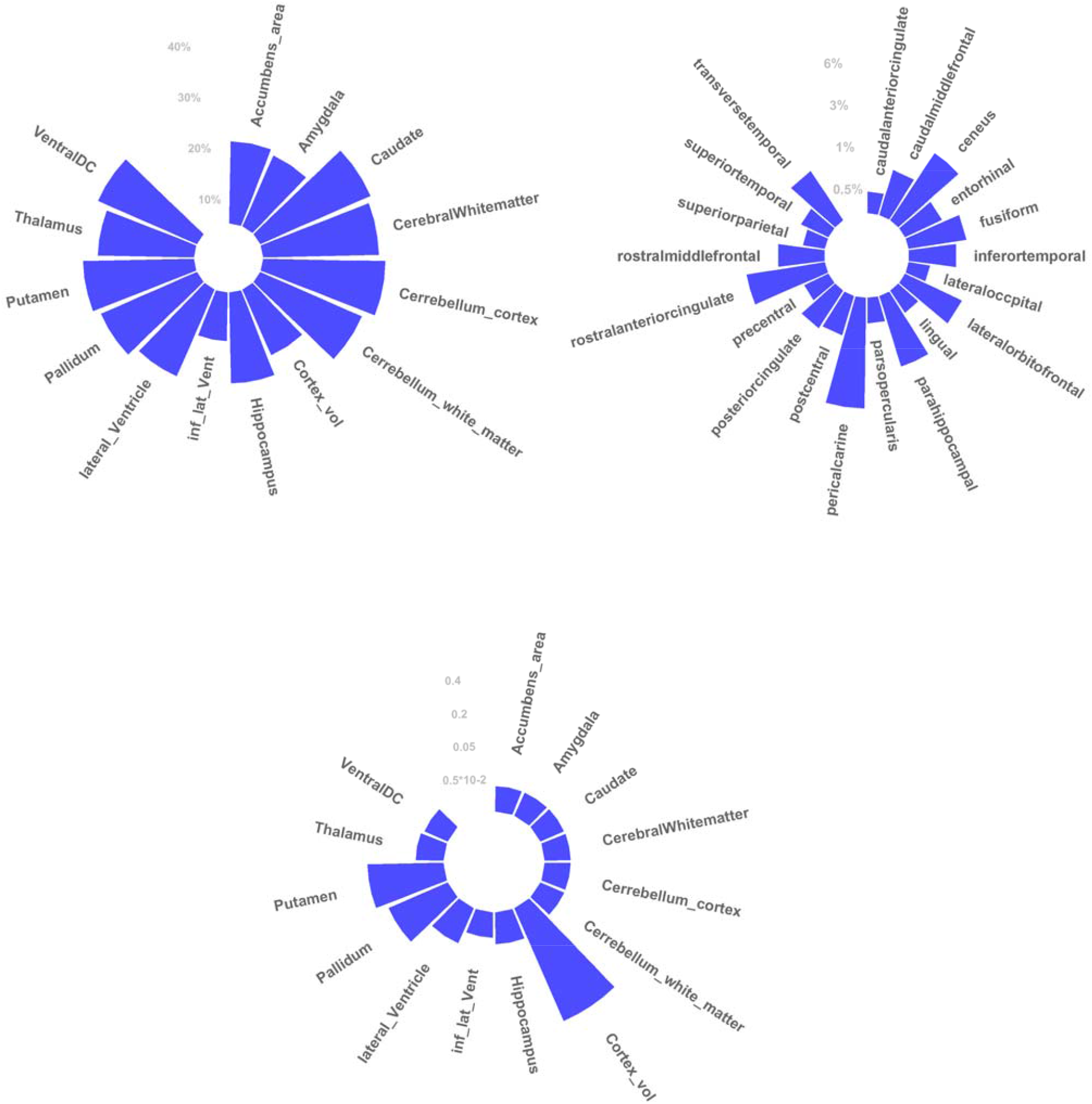
Circosplot of left hemisphere of the Subcortical brain volume PIDS. Figure 4.1 circosplot of heritability, Figure 4.2 circosplot of variance explained, and Figure 4.3 circosplot of P-thresholds.

**Figure 5.**
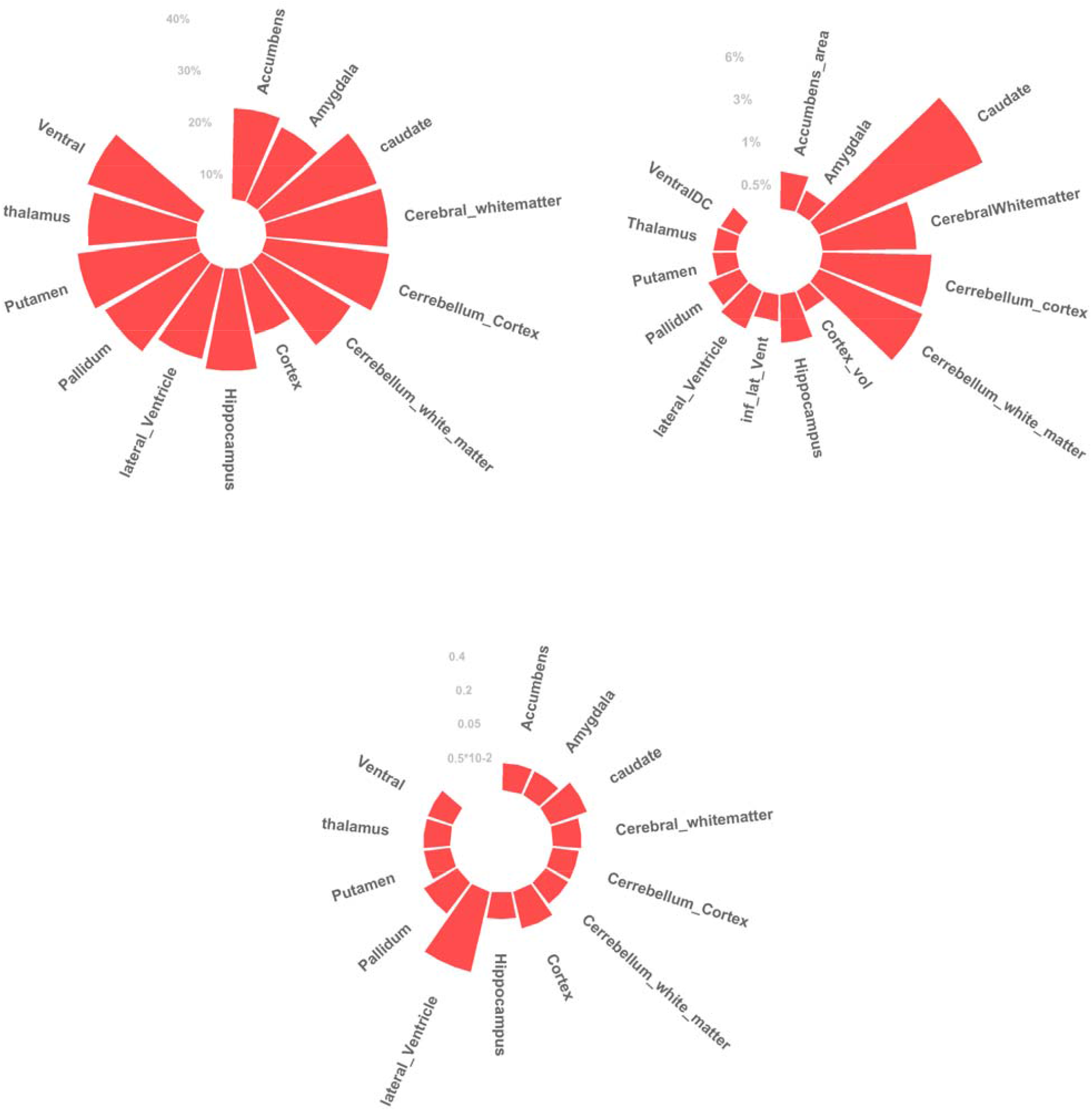
Circosplot of right hemisphere of the Subcortical brain volume PIDS. Figure 5.1 circosplot of heritability, Figure 5.2 circosplot of variance explained, and Figure 5.3 circosplot of P-thresholds.

Amongst PIDS scoring highest in genetic effect size were global measures cerebellum white matter, cerebellum cortex, and cerebral white matter, consistently found across hemispheres. Amongst the PIDS with the lowest variance explained > 1%, were multiple gyral volumes, superior and middle temporal volume, isthmus cingulate gyrus and the global measure cortex volume, consistently found across hemispheres (supplementary table 3). The Subcortical and Cortical volumes used for the PIDS differ significantly in heritability. Heritability was found to be a good predictor for the genetic effect size of the PIDS (F (1,70) = 11.77, *p = 0.001*) with an *R*^2^ of 0.132. Moreover, with the greater heritability found in the Subcortical volumes used for the PIDS we would expect the Subcortical PIDS to outperform the cortical PIDS. However, this was not the case with no significant differences found between Cortical and Subcortical PIDS in genetic effect size, P-threshold, or number of independent markers. The PIDS are consistent in their performance across the four different subgroups. Interestingly, small correlations (−0.4 < *h*^*2*^ < 0.4) were found for genetic effect size, P-threshold, and number of SNPS between hemispheres for both cortical and subcortical PIDS. A sizeable correlation (*r* = 0.77) was found between the cortical hemispheres for heritability. 22 PIDS out of the original 94 that were excluded from further analyses after stage 1. This exclusion could potentially have influenced the correlation values we observe in the analysis.

Most PIDS (52 out of 72) had an optimal P-threshold between 0.001 and 1, reliably reflecting the polygenic nature of IDP’s. However, the 10 PIDSs with the lowest P-thresholds ranged between 0.00005 and 0.00000005, conflicting with the expected polygenicity. The PRSice generated bar plots further illustrate these findings (supplementary figure 2.1-2.80).

The PIDS amongst the lowest ranging P-thresholds produce bar plots with inconsistent R-squared values across different P-thresholds. This may indicate overfitting of these specific PIDS. The bar plots for 28 PIDSs were of note due to consistent performance across most p-thresholds. Amongst these 28 PIDS were the global measures: cerebellum cortex, cerebellum white matter, cerebral white matter, and cortex volume, consistently found across both hemispheres. PIDS with consistent performance across a range of p-thresholds also performed optimally with inclusive P-thresholds (0.01-1) (supplementary table and figure’s 3.1-3.80).

### PIDS and ADHD status

In stage two of the project, PIDS were used to predict ADHD diagnosis and subthreshold ADHD using linear mixed modeling. Results indicated six out of the 72 PRS to be nominally associated with ADHD diagnosis of which none remained after FDR correction (table 2) (*p ,0.05)*. These 6 PIDS were approximately normally distributed (supplementary figure 1). To account for the large number of tests across 94 IDP’s, we considered a false-discovery rate (FDR) of 0.05. After FDR correction for the 72 performed tests, no significant association remained. No interaction was found with sex, site, or batch for any of the PIDS. The six PIDS were used to predict ADHD persistence using linear regression models. No significant relationship was found between any of the six IDP PRS and ADHD persistence (all *P* > 0.4). Nor was any significant relationship found for partial remission or remission category. The six PIDS were also used to predict determinants of long-term ADHD (DELTA). No significant effects were found for DIVA interview in DELTA diagnosis based on the DSM5, diagnosis based on DSM4, updated persistence and updated partial remittance.

**Table 2.** The five MRI traits showing promise in predicting ADHD diagnosis. Rh = right hemisphere lh = left hemisphere

| Phenotype | P threshold | Coefficient | Standard error | P | SNPs | FDR corrected P |
| --- | --- | --- | --- | --- | --- | --- |
| rh isthmuscingulate volume | 0.0233 | 0.265 | 1587.95 | 0.0459 | 36501 | 0.612 |
| rh postcentral_volume | 0.0221 | -0.391 | 1542.76 | 0.0182 | 36051 | 0.512 |
| Lh_Lateroccipital_volume | 0.05 | 0.255 | 0.1852 | 0.0268 | 69543 | 0.545 |
| Lh_posterior_Cingulate_volume | 0.01 | -2.148 | 1.0877 | 0.0483 | 17681 | 0.612 |
| Lh_Ventral_DC | 0.001 | 0.812 | 0.3712 | 0.0287 | 3090 | 0.524 |
| Rh_Ventral_DC | 0.001 | 0.949 | 0.3504 | 0.0068 | 3096 | 0.495 |

### Power analysis

Polygenic scores were constructed associating IDPS with ADHD based on a panel of 100,000 independent markers with a discovery sample of 30,000 and an effective target sample of 964. Assuming prevalence of 4%, a SNP-*h*^*2*^ of 22% and a genetic covariance of 6% we achieve 20% power. Due to the small genetic covariance between the IDPS and ADHD a larger sample size is required to achieve acceptable power. However, when increasing both target and discovery sample sizes, leaps in power can be achieved. When increasing the discovery sample to 50,000 and target sample to 3,000, 0.80 power would be the result (figure 6). The sample sizes required for this amount of power are an order of magnitude higher than currently available but are achievable in future research. The UKB sample is projected to increase to a GWAS imaging sample of 100,000 participants. Moreover, with large increases in the GWAS discovery sample, larger SNP-*h*^*2*^ closer to the narrow-sense heritability can be achieved. This increase in genetic effect size was not considered for the estimated increase in power, leaving this estimation to be somewhat conservative.

**Figure 6.**
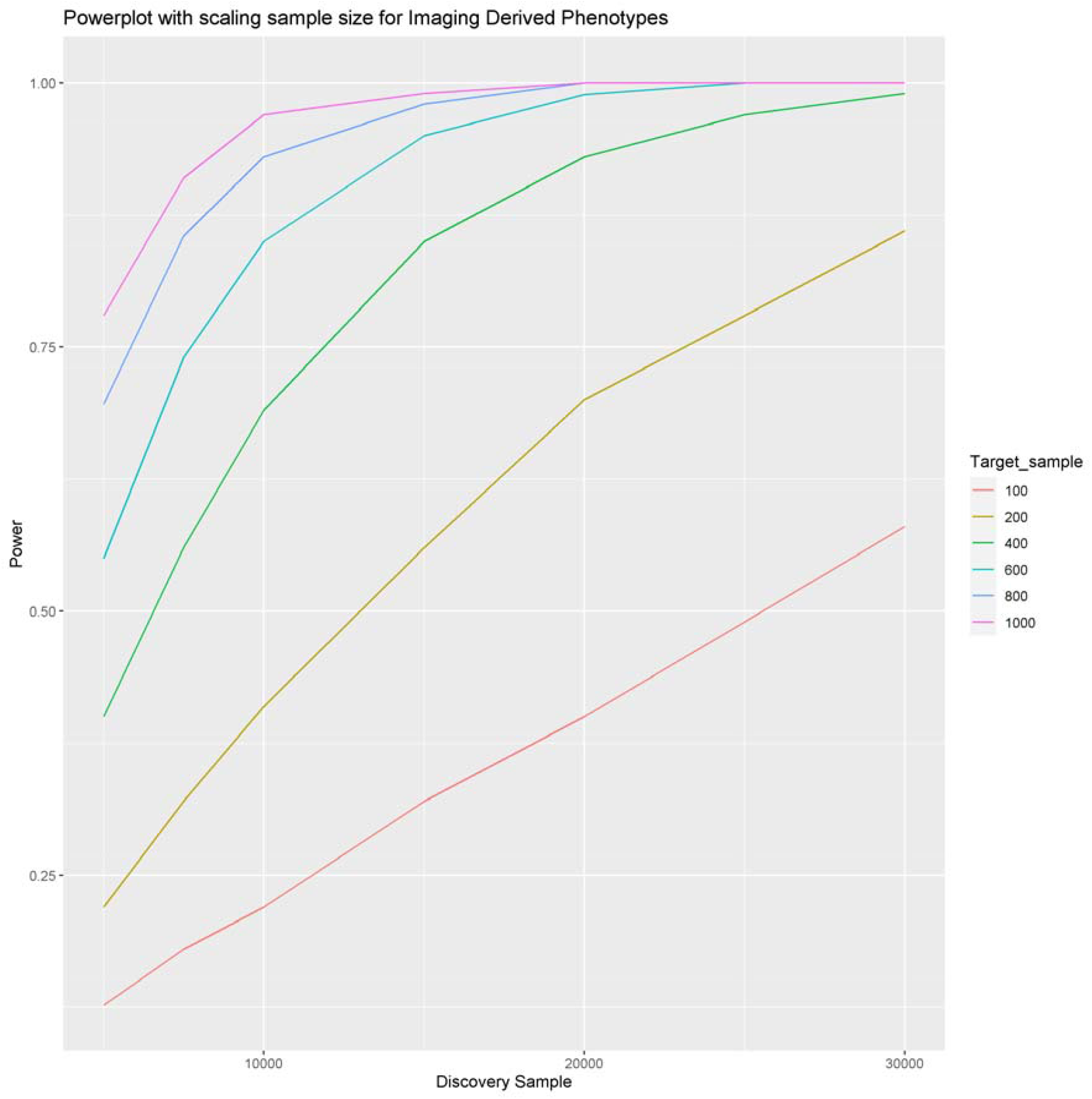
Powerplot scaling with sample size for the PIDS sample. Starting point of the plot shows the PIDS achieved 0.96 power with a discovery sample of ≈ 30.000 and a target sample of ≈ 1000. However, a satisfactory 0.80 power can also be achieved with a discovery sample of 30000 and a target sample of 200 For each sample size the P-threshold is applied that leads to the highest power.

**Figure 7.**
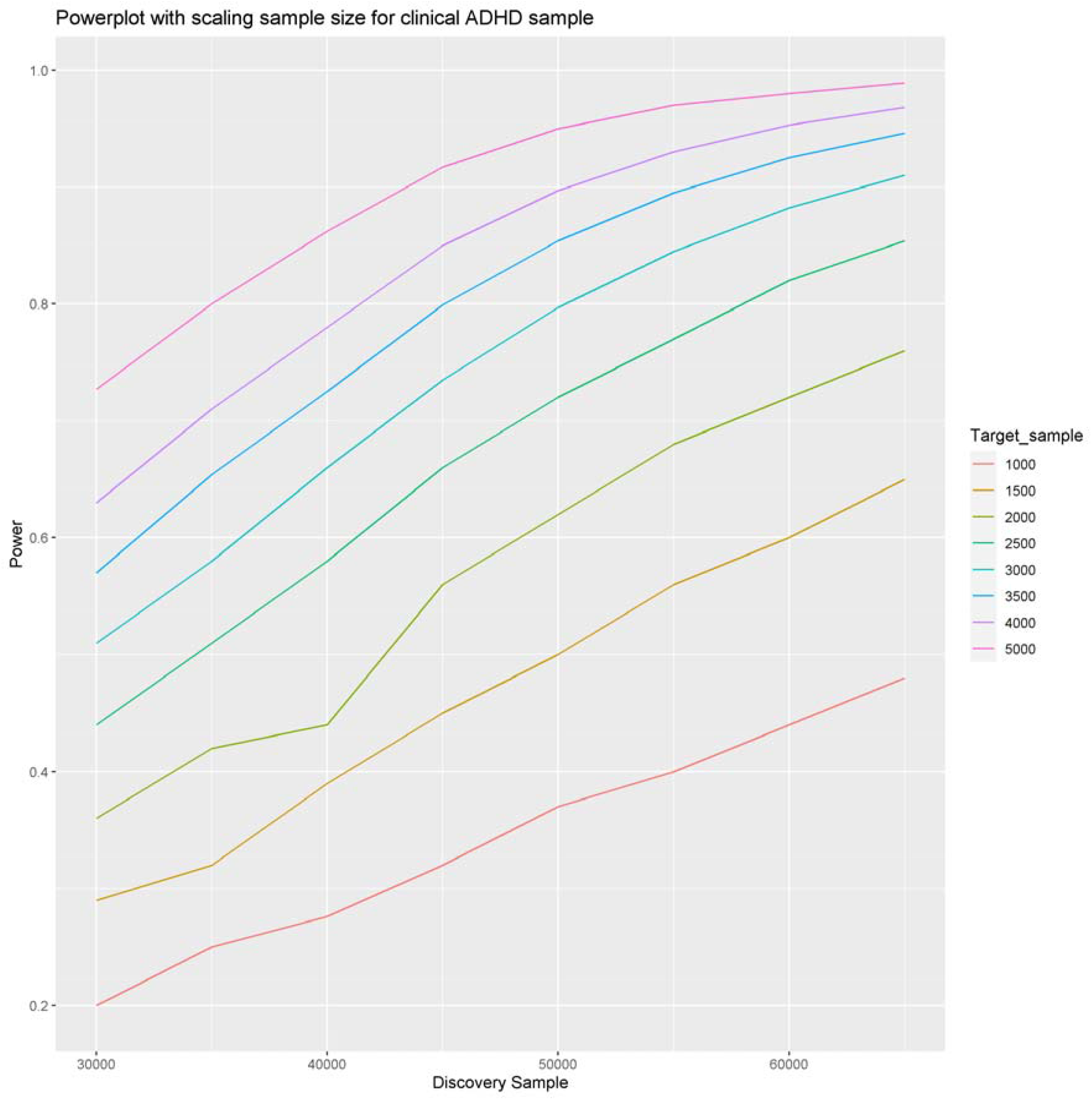
Powerplot scaling with sample size for the clinical ADHD sample. Starting point of the plot shows the achieved 0.20 power with a discovery sample of ≈ 30.000 and a target sample of ≈ 1000. A satisfactory 0.80 power can be achieved with a discovery sample of 50000 and a target sample of 3000. For each sample size the P-threshold is applied that leads to the highest power.

## Discussion

This study investigated the relationship between the combined effect of imaging derived phenotype associated SNPs and ADHD-related traits by using polygenic imaging derived scores or PIDS. Where previous work tended to use clinically derived PRS to predict neuroimaging traits, we use IDPs to create PIDS and predict ADHD-related traits. We showed PIDS can be successfully derived with the majority robustly reflecting the genetically driven inter-individual differences in brain structure. However, in the ADHD sample, the PIDS did not describe inter-individual differences in related clinical variables. This lack of performance is in part due to insufficient power to detect the small sub portion of genetic variation shared between ADHD and the IDPs.

The results indicate that the majority of PIDS were predictive of their respective IDP in an independent target sample. The polygenic nature of the IDPs was reflected through consistent performance of most cortical and sub-cortical PIDS across a wide variety of P-value thresholds (supplementary figures 2.1-1.80 & 3.1-2.80). PRSice-generated bar plots and high-resolution plots reflect most PIDS to be stable predictors of out-of-sample cortical and subcortical brain structure despite the large age difference in the base (UKB) and target (NeuroIMAGE) datasets. This large degree of predictive PIDS indicates GWAS data such as the UKB can be successfully used to predict inter-individual variance. Moreover, the most predictive PIDS reached variance explained ranges upwards of 3% - 5.7%. A trend of association was found between the right hemisphere postcentral volume, left hemisphere thalamus, left hemisphere putamen, and both hemispheres of the isthmus cingulate volume PIDS and ADHD. The variance in the ADHD phenotype explained by the PIDS did not reach beyond 1%. None of these associations survived FDR correction. No associations were found between PIDS and ADHD persistence-remittance variables. In short, we did not find any significant association between PIDS and ADHD.

The SNP-heritability of cortical volumes being greater than subcortical volumes was expected to result in greater performance of the cortical PIDS in genetic effect size, significance, and power. Interestingly, no differences were found in genetic effect size, nor significance. Moreover, low correlations were found between hemispheres for genetic effect size, heritability, and p-thresholds. However, Cortical PIDS did achieve greater power than Subcortical PIDS.

The results of this project should be interpreted considering some limitations. First, The effects sizes for brain structure in association with ADHD are generally very small (Hoogman et al al., 2019). Due to the small genetic covariance between ADHD and the PIDS our target sample was insufficiently large to meaningfully inter-individual differences in related clinical variables. We reached 0.20 power with the current sample of 30,000 participants in the GWAS dataset and 744 in the target sample. Clinical sample size remains a big issue in neuroimaging research. However, we project with an increase to 50.000 participants in the UKB GWAS and 3000 in the clinical NeuroIMAGE target sample we will achieve 0.80 power. Interesting patterns/trends emerged from the analysis, with this increase in target and base datasets these should be pursued further.

Second, genetic effects on brain measures are inherently correlated (fleary et al, 2021; Hofer et al., 2020; Smith et al., 2021). with global measures being the most heritable. We did not use global brain measures as covariates in calculating the PIDS. The UKB GWAS has already been corrected for total head volume accounting for a degree of the correlation between the global brain measures and our PIDS. If we were to remove variation by accounting for global brain measures our ability to quantify the SNP-effects associated with IDPs would decrease substantially.

PIDS may have use for studying psychiatric disorders in the future, as a basis for better understanding underlying biology in addition to heterogeneous diagnosis (Whalley et al., 2012) The application of PIDS extend beyond single disorders into transdiagnostic vulnerability, researching psychiatric disorders starting from the brain. Focusing on imaging derived traits rather than syndromes and using the resultant knowledge and statistics as a modelling basis for bottom-up clinical research, represents a framework shift for intermediate phenotype use in psychiatric genetics. Disorders that are phenotypically dissimilar may still share common neural vulnerability (Beauchaine & Constantino, 2017). Hence, we chose to start with the definition of this neural vulnerability and asses the genetic component to this trait rather than that of a disorder. These transdiagnostic and data-driven applications of intermediate phenotypes can be useful if we are to evaluate neural vulnerabilities and their interactions with or role in genetic risk at the root of behavior.

This project presents a new approach to characterize the genetic underpinnings of psychiatric traits by quantifying the common variants involved in relevant brain variation and using them to profile individual patients. GWAS data can be used successfully to explain Individual variance. However, this same success has yet to be achieved for variance in ADHD. Currently, the implementation PRS is hindered by weaker application to non-European ancestry and limited translation of percentile loadings to lifetime risk. With improved diverse ancestry databases and a combining of environmental and clinical risk factors with the PRS, polygenic scores based on imaging derived phenotypes could serve as a more objective tool for investigating underlying neurobiology and genetic architecture complementary to syndromic classification. Ultimately, PIDS may have potential value when included in prediction and classification models. Profiling individuals in terms of genetic influences on different areas of the brain could for example be useful when describing heterogeneity in etiological factors, prognosis, or treatment response. These findings encourage future studies to use PIDS to translate fundamental neuroimaging genetics to the field of psychiatry and beyond.

## Data Availability

All data produced in the present study are available upon reasonable request to the authors

https://open.win.ox.ac.uk/ukbiobank/big40/

## Acknowledgements

We are appreciative to UK Biobank for making the data available and to all UK Biobank participants who donated their time and made this resource possible. The BIG40 Open Data Server is provided by the Wellcome Centre for integrative Neuroimaging, which is supported by center funding from the Wellcome Trust (203139/Z/16/Z). Compute resources were provided by the Oxford Biomedical Research Computing (BMRC) facility (a joint development between Oxford’s Wellcome Centre for Human genetics and Big Data Institute, supported by Health Data Research UK and the NIHR Oxford Biomedical Research Centre). Furthermore, we would like to thank all participants and families who participated in the IMAGE, NeuroIMAGE, and DELTA studies. The DELTA project was funded by a Radboudumc Hypatia Fellowship (E. S.). NeuroIMAGE was supported by a Dutch Research Council (NWO) Large Investment Grant (no. 1750102007010) and NWO Brain & Cognition an Integrative Approach Grant (no. 433–09-242 to J.K.B.), and grants from Radboud University Medical Center, University Medical Center Groningen and Accare, and VU University Amsterdam. B.F. has received educational speaking fees from Medice. J.K.B. has been in the past 3 years a consultant to / member of advisory board of / and/or speaker for Takeda/Shire, Roche, Medice, Angelini, Janssen, and Servier. He is not an employee of any of these companies, and not a stock shareholder of any of these companies. All other authors report no biomedical financial interests or potential conflicts of interest.

## Literature

Beauchaine, T. P., & Constantino, J. N. (2017). Redefining the endophenotype concept to accommodate transdiagnostic vulnerabilities and etiological complexity. 11, 769–780.

Bigos, K. L., & Weinberger, D. R. (2010). Imaging genetics-days of future past. NeuroImage, 53(3), 804–809. https://doi.org/10.1016/j.neuroimage.2010.01.035

Bonvicini, C., Faraone, S. V., & Scassellati, C. (2016). Attention-deficit hyperactivity disorder in adults: A systematic review and meta-analysis of genetic, pharmacogenetic and biochemical studies. Molecular Psychiatry, 21(7), 872–884. https://doi.org/10.1038/mp.2016.74

Bralten, J., Greven, C. U., Franke, B., Mennes, M., Zwiers, M. P., Rommelse, N. N. J., Hartman, C., van der Meer, D., O’Dwyer, L., Oosterlaan, J., Hoekstra, P. J., Heslenfeld, D., Arias-Vasquez, A., & Buitelaar, J. K. (2016). Voxel-based morphometry analysis reveals frontal brain differences in participants with ADHD and their unaffected siblings. Journal of Psychiatry and Neuroscience, 41(4), 272–279. https://doi.org/10.1503/jpn.140377

Choi, S. W., Mak, T. S. H., & O’Reilly, P. F. (2020). Tutorial: a guide to performing polygenic risk score analyses. Nature Protocols, 15(9), 2759–2772. https://doi.org/10.1038/s41596-020-0353-1

Choi, S. W., & Reilly, P. F. O. (2019). PRSice-2□: Polygenic Risk Score software for biobank-scale data. November 2018, 1–6. https://doi.org/10.1093/gigascience/giz082

Ck, C., Sitarenios, G., Jd, P., Jn, E., Parent, C., Conners, C. K., Sitarenios, G., Parker, J. D. A., & Epstein, J. N. (1998). The Revised Conners ‘ Parent Rating Scale (CPRS-R): Factor Structure, Reliability, and Criterion Validity. April 2014. https://doi.org/10.1023/A

Conners, C. K., Wells, K. C., Parker, J. D. A., Sitarenios, G., John, M., & Powell, J. W. (1998). A New Self-Report Scale for Assessment of Adolescent Psychopathology□: A New Self-Report Scale for Assessment of Adolescent Psychopathology□: Factor Structure, Reliability, Validity, and Diagnostic Sensitivity. January. https://doi.org/10.1023/A

Demontis, D., Walters, G. B., Athanasiadis, G., Walters, R., Gudmundsson, O. O., Magnusson, S. H., & Baldursson, G. (2022). Genome-wide analyses of ADHD identify 27 risk loci, refine the genetic architecture and implicate several cognitive domains. 1–40.

Desikan, R. S., Ségonne, F., Fischl, B., Quinn, B. T., Dickerson, B. C., Blacker, D., Buckner, R. L., Dale, A. M., Maguire, R. P., Hyman, B. T., Albert, M. S., & Killiany, R. J. (2006). An automated labeling system for subdividing the human cerebral cortex on MRI scans into gyral based regions of interest. NeuroImage, 31(3), 968–980. https://doi.org/10.1016/j.neuroimage.2006.01.021

Destrieux, C., Fischl, B., Dale, A., & Halgren, E. (2010). Automatic parcellation of human cortical gyri and sulci using standard anatomical nomenclature. NeuroImage, 53(1), 1–15. https://doi.org/10.1016/j.neuroimage.2010.06.010

Dudbridge, F. (2013). Power and Predictive Accuracy of Polygenic Risk Scores. 9(3). https://doi.org/10.1371/journal.pgen.1003348

Euesden, J., Lewis, C. M., & Reilly, P. F. O. (2016). PRSice□: Polygenic Risk Score software. 1–24.

fleary et al. (2021). 乳鼠心肌提取 HHS Public Access. Physiology & Behavior, 176(3), 139–148. https://doi.org/10.1126/science.aay6690.The

Hofer, E., Roshchupkin, G. V., Adams, H. H. H., Knol, M. J., Lin, H., Li, S., Zare, H., Ahmad, S., Armstrong, N. J., Satizabal, C. L., Bernard, M., Bis, J. C., Gillespie, N. A., Luciano, M., Mishra, A., Scholz, M., Teumer, A., Xia, R., Jian, X., … Zhou, J. (2020). Genetic correlations and genome-wide associations of cortical structure in general population samples of 22,824 adults. Nature Communications, 11(1). https://doi.org/10.1038/s41467-020-18367-y

Krain, A. L., & Castellanos, F. X. (2006). Brain development and ADHD. Clinical Psychology Review, 26(4), 433–444. https://doi.org/10.1016/j.cpr.2006.01.005

Kuntsi, J., Neale, B. M., Chen, W., Faraone, S. V, & Asherson, P. (2006). Behavioral and Brain Functions The IMAGE project□: methodological issues for the molecular genetic analysis of ADHD. 13, 1–13. https://doi.org/10.1186/1744-9081-2-27

Liu, S., Smit, D. J. A., Abdellaoui, A., Wingen, G. A. Van, & Verweij, K. J. H. (2021). Brain structure and function show distinct relations with genetic.

Medland, S. E., Shumskaya, E., Jahanshad, N., Zeeuw, P. De, Szekely, E., Sudre, G., Wolfers, T., Onnink, A. M. H., Dammers, J. T., Mostert, J. C., Vives-gilabert, Y., Kohls, G., Oberwelland, E., Seitz, J., Schulte-rüther, M., Ambrosino, S., Doyle, A. E., Høvik, M. F., Dramsdahl, M., … Franke, B. (2017). Subcortical brain volume differences in participants with attention deficit hyperactivity disorder in children and adults□: a cross-sectional mega-analysis. 4(April), 310–319. https://doi.org/10.1016/S2215-0366(17)30049-4

Müller, U. C., Asherson, P., Banaschewski, T., Buitelaar, J. K., Ebstein, R. P., & Eisenberg, J. (2011). The impact of study design and diagnostic approach in a large multi-centre ADHD study. Part 1□: ADHD symptom patterns The impact of study design and diagnostic approach in a large multi-centre ADHD study. Part 1□: ADHD symptom patterns. 54(April).

Parker, J. D. A., & Sitarenios, G. (1999). Self-ratings of ADHD symptoms in adults I□: Factor structure and normative data. October. https://doi.org/10.1177/108705479900300303

Romme, I. A. C., de Reus, M. A., Ophoff, R. A., Kahn, R. S., & van den Heuvel, M. P. (2017). Connectome Disconnectivity and Cortical Gene Expression in Patients With Schizophrenia. Biological Psychiatry, 81(6), 495–502. https://doi.org/10.1016/j.biopsych.2016.07.012

Ronald, A., de Bode, N., & Polderman, T. J. C. (2021). Systematic Review: How the Attention-Deficit/Hyperactivity Disorder Polygenic Risk Score Adds to Our Understanding of ADHD and Associated Traits. Journal of the American Academy of Child and Adolescent Psychiatry. https://doi.org/10.1016/j.jaac.2021.01.019

Smith, S. M., Douaud, G., Chen, W., Hanayik, T., Alfaro-Almagro, F., Sharp, K., & Elliott, L. T. (2021). An expanded set of genome-wide association studies of brain imaging phenotypes in UK Biobank. Nature Neuroscience, 24(MaY). https://doi.org/10.1038/s41593-021-00826-4

Watanabe, K., Stringer, S., Frei, O., Umicevic Mirkov, M., de Leeuw, C., Polderman, T. J. C., van der Sluis, S., Andreassen, O. A., Neale, B. M., & Posthuma, D. (2019). A global overview of pleiotropy and genetic architecture in complex traits. Nature Genetics, 51(9), 1339–1348. https://doi.org/10.1038/s41588-019-0481-0

Weinberger, R. (2011). NIH Public Access. 79(1), 9–22. https://doi.org/10.1016/j.biopsycho.2008.03.015.Neurobiology

Whalley, H. C., Papmeyer, M., Sprooten, E., Romaniuk, L., Blackwood, D. H., Glahn, D. C., Hall, J., Lawrie, S. M., Sussmann, J. E., & McIntosh, A. M. (2012). The influence of polygenic risk for bipolar disorder on neural activation assessed using fMRI. Translational Psychiatry, 2(June), 1–8. https://doi.org/10.1038/tp.2012.60

Yang, J., Finucane, H. K., Gusev, A., Ripke, S., Genovese, G., Loh, P., Bhatia, G., Lindstro, S., Do, R., Zaitlen, N., Pasaniuc, B., Belbin, G., Kenny, E. E., Schierup, M. H., Purcell, S., Chasman, D., Neale, B., & Goddard, M. (2015). Modeling Linkage Disequilibrium Increases Accuracy of Polygenic Risk Scores. 576–592. https://doi.org/10.1016/j.ajhg.2015.09.001

Yang, J., Yan, B., Zhao, B., Fan, Y., He, X., Yang, L., Ma, Q., Zheng, J., Wang, W., Bai, L., Zhu, F., & Ma, X. (2020). Assessing the Causal Effects of Human Serum Metabolites on 5 Major Psychiatric Disorders. Schizophrenia Bulletin, 1–10. https://doi.org/10.1093/schbul/sbz138

